# Distinct biochemical phenotypes of HIV exposed infants driven by antiviral medication

**DOI:** 10.64898/2026.01.28.26344948

**Authors:** Shujian Zheng, Jasmine Chong, Jennifer Canniff, Michael J Johnson, Maheshwor Thapa, Elizabeth Aiken, Shabir Madhi, Adriana Weinberg, Shuzhao Li

## Abstract

Pregnant women with HIV control viral replication with antiretrovirals and give birth to HIV-exposed uninfected infants (HEU). The children, however, exhibit increased morbidity and mortality due to severe infections, as well as cognitive and growth abnormalities. In this study, we performed high-resolution, untargeted metabolomics on 123 HIV-exposed mother-baby pairs and 117 control pairs without HIV. High concentrations of the antiretroviral efavirenz and its metabolites were detected in maternal blood and cord blood. The metabolomic differences between HEU participants and controls reflect perturbed pathways of steroids, tryptophan and bile acids, and they largely consisted of metabolites that were correlated with efavirenz concentrations within the HEU group. The results suggest a major contribution of the drug to the abnormal biochemical profile of HEU infants born to mothers treated with efavirenz.

---

Epidemiological evidence shows that in utero exposures to infections and chemical substances, including therapeutics, have profound impacts on population health[1-5]. A well-studied area is children born to mothers with HIV, who are not infected by HIV (HIV-Exposed Uninfected or HEU). The HEU children are known to exhibit increased morbidity and mortality due to severe infections, as well as cognitive and growth abnormalities when compared to HIV-unexposed children (HIV-Unexposed Uninfected or HUU)[6-9]. This poses a significant health burden with over 16 million affected children in 2023, most of whom live in sub-Saharan Africa [9]. The extent to which exposure to antiretrovirals contributes to undesirable health outcomes is incompletely understood.

Untargeted metabolomics has recently become a promising approach by quantifying the exposures and biological effects simultaneously[10-13]. Previous metabolomic studies in HEU infants were technologically limited and measured small numbers of metabolites/lipids in small cohorts [14-17]. In this study, we performed untargeted, high-resolution metabolomics and lipidomics in 123 HEU mother-infant pairs and 117 HUU control pairs. The demographic characteristics of the study population was previously described[18]. Notably, all mothers with HIV received antiretroviral therapy during pregnancy, including efavirenz in 92% of them. The metabolomic analysis was performed with hydrophilic interaction chromatography (HILIC), and lipidomics reverse phase (RP) chromatography, both using positive and negative electrospray ionization (ESI+/-, see Methods). The data were generated on plasma samples from maternal peripheral blood collected at delivery and cord blood. Unsupervised principal component analysis (PCA) showed clear separations of the mothers and infants by both sampling method and by HIV status (Fig. 1a, Suppl. Fig. 1a). The difference was driven by large numbers of metabolites with significantly different concentrations (FDR < 0.05, 658 RP ESI-features Fig. 1b; 1827 HILIC ESI-features, Suppl. Fig. 1b).

**Figure 1.**
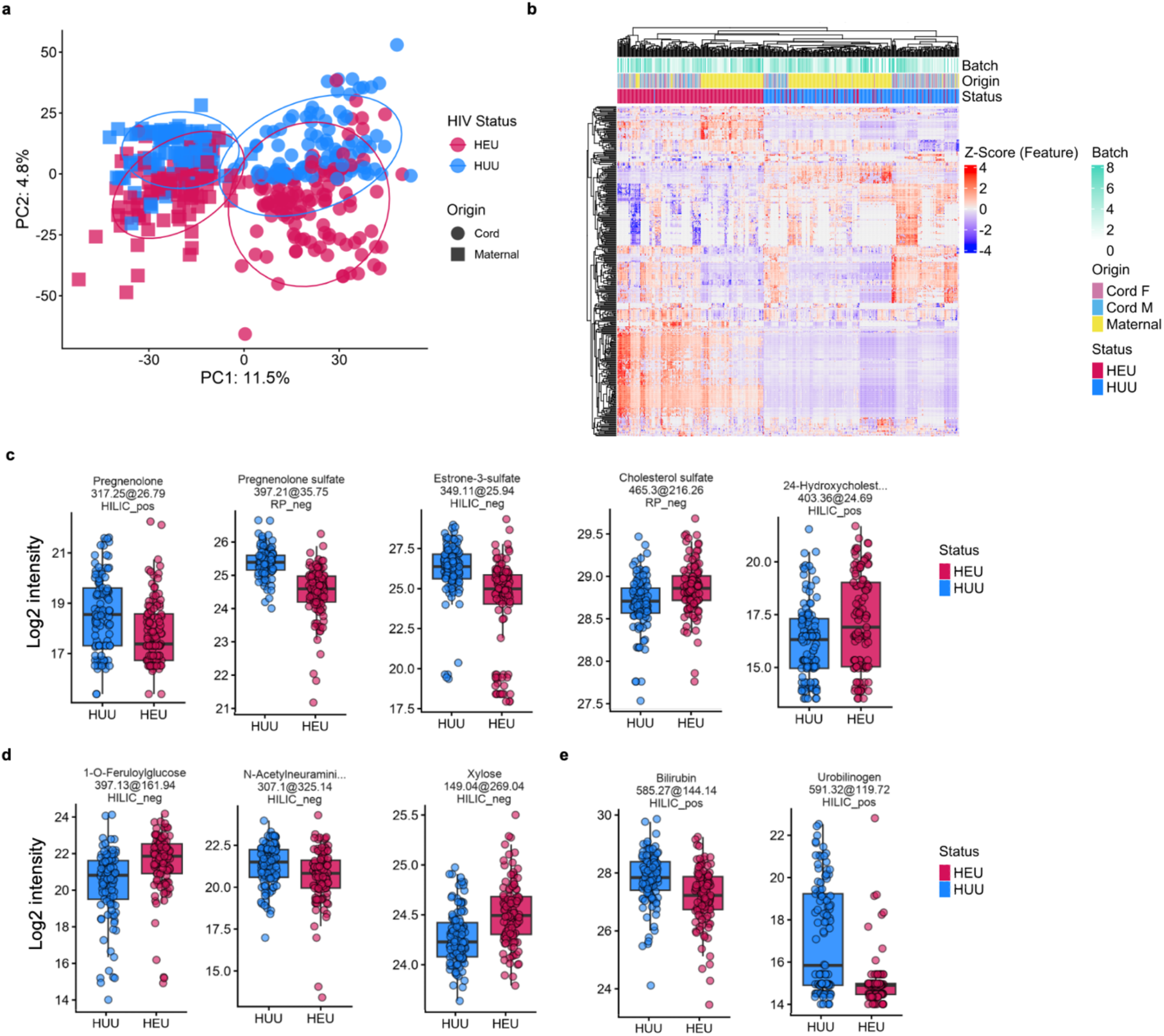
Distinct biochemical phenotypes of mother-infant dyads by metabolomic profiling. a) PCA plot showing the separation of HEU/HUU and maternal/cord plasma samples in metabolomics, using all features in RP-(negative ionization from reverse phase chromatography). b) Cluster heatmap of metabolomic features (q<0.05, RP-), samples largely cluster by HIV exposure status. c-e) Boxplots of example significant metabolites.

HEU cord blood displayed altered steroid metabolism, which included lower pregnenolone, pregnenolone sulfate, and estrone sulfate, but higher cholesterol derivatives (Fig. 1c). Notably, many sulfated metabolites were significant in the result, possibly due to the involvement of sulfotransferases in drug metabolism [19]. Disruptions of sugar and bilirubin metabolism were also detected (Fig. 1d,e; Suppl Fig. 2a,b). Tryptophan and its metabolites, such as kynurenic acid, show different direction of alteration in HEU and HUU infants (Suppl Fig. 2c). Differentially abundant bile acids due to HIV exposure include glycocholic and taurochenodesoxycholic acid (Suppl Fig. 2d). Multiple polyunsaturated fatty acids (PUFAs), commonly linked to CYP2B6 activity, were perturbed (Suppl Fig. 2e). Given the critical roles of pregnenolone and PUFAs in fetal development[20, 21], this suggests a mechanism for impaired growth in HEU infants. The dysregulation of bile acids and tryptophan metabolisms may play a role in their impaired immune and neural developments[22-24].

Differences in steroids and estrogens were more significant in the mothers than infants, including reductions in DHEA-S, estrone sulfate, glucuronidated cholesterol and sulfated steroids (Suppl. Fig. 2f,g). Metabolites with significantly different concentrations in HEU and HUU were shared between mothers and infants. Additional examples included higher bilirubin derivatives in HUU, elevated taurocholic and glycocholic acid in HEU, decreased PAGln and 3-methoxyphenol sulfate, and elevated dopamine 3-0 sulfate in HEU (Suppl. Fig. 2h). Network analysis using the mummichog software[25] highlighted activities in steroid metabolism in both HEU mothers and infants (Suppl. Fig. 3).

Among the top differential features by HEU status were efavirenz and its metabolites (Fig. 2a). The chemical identity of efavirenz and its metabolite 8-hydroxyefavirenz was confirmed by matched mass-to-charge ratio (m/z), retention time and MS/MS spectra to authentic standards (Fig. 2b). The drug was detected in the cord blood of most HEU infants, and strong correlations were observed between efavirenz and 8-hydroxyefavirenz (Fig. 2c). Similar patterns were observed in mothers with HIV (Fig. 2d,e; Suppl. Fig. 4a). These data are consistent with previous studies demonstrating that the drug readily crosses the placenta and exposes the fetuses at high doses[26, 27].

**Figure 2.**
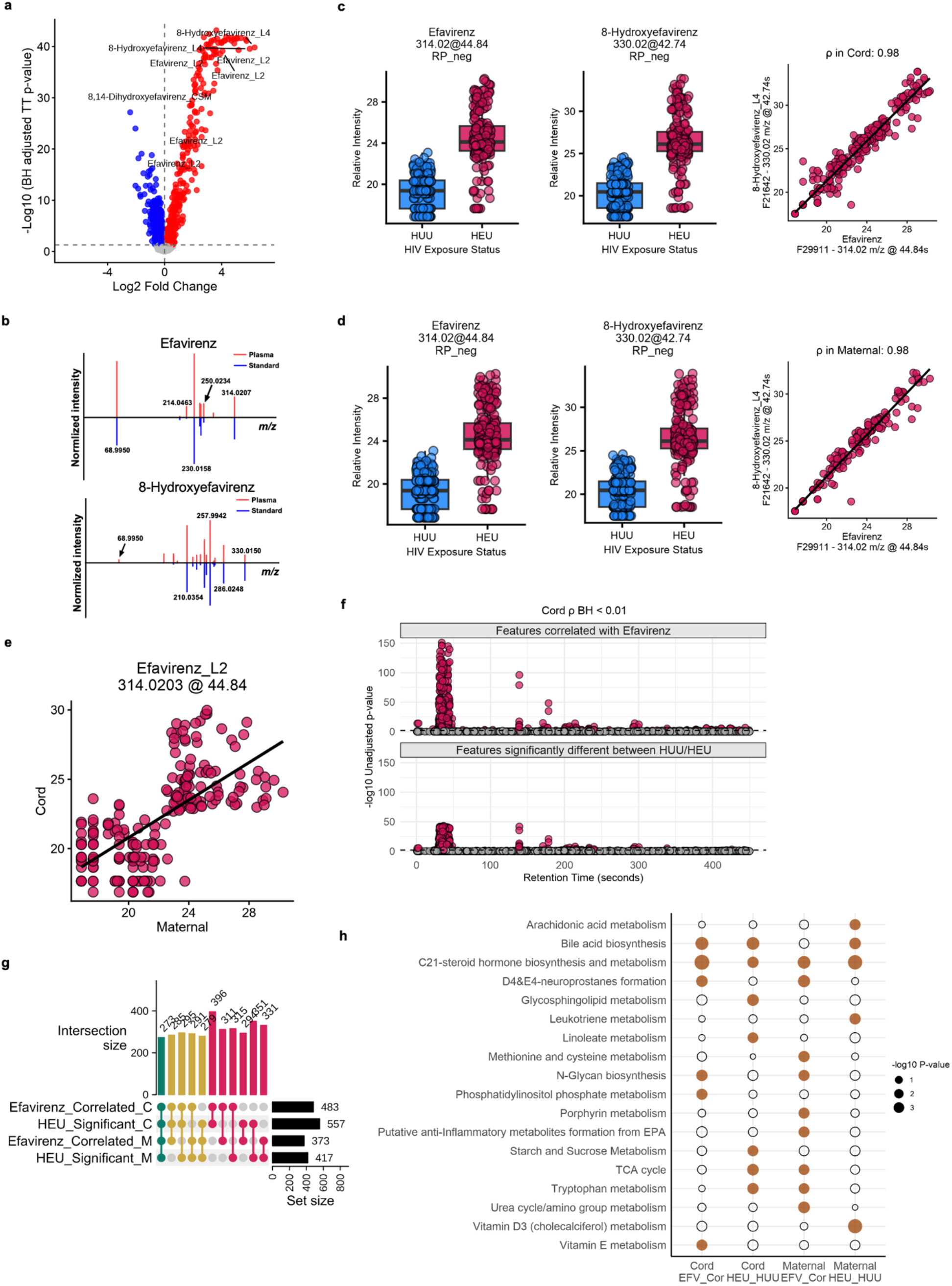
Efavirenz drives differential metabolites in HEU infants. a) Volcano plot of metabolomic features significantly different between HEU and HUU infants (RP-, Efavirenz and its metabolites are labeled). b) MS/MS identification of efavirenz and 8-hydroxyefavirenz. c) Detection of efavirenz and 8-hydroxyefavirenz in infants (RP-); each dot in boxplot represents a subject. The baselines were not zero because the compounds have residuals in chromatographic columns. d) Detection of efavirenz and 8-hydroxyefavirenz in mothers (RP-); each dot in boxplot represents a subject. e) Correlation of efavirenz levels between infants and their mothers. f) Manhattan plot comparing the distribution of features correlated with Efavirenz versus features that are significantly different due to HIV exposure in cord plasma samples along the retention-time axis (RP-). g) UpSet plot showing the intersection of features significantly correlated with efavirenz and features significantly associated with HIV exposure status (RP-). h) Significant pathways overlap between efavirenz correlates and HEU/HUU differences (HILIC+). The HILIC+ data have more pathway coverage compared to RP- data (Suppl Fig. 4c).

To further understand the role of efavirenz in the metabolic alterations of HEU mother-infant pairs, we carried out a metabolome-wide association study (MWAS), taking advantage of the broad range of efavirenz concentrations in HEU infants and their mothers. In the HEU infants, 483 features were significantly correlated with efavirenz (Pearson’s correlation, FDR < 0.01), as visualized on a Manhattan plot along LC elution time (Fig. 2f, top). Compared to the 557 features significantly different between HEU and HUU (Fig. 2f, bottom), the major feature clusters appeared strikingly similar. The similarity extended to the maternal data (Suppl Fig. 4b). Indeed, comparisons between the four groups, efavirenz MWAS and HEU/HUU differences in infants and mothers, show that the significant features largely overlapped (Fig. 2g). The top pathways enriched in efavirenz-correlated metabolites also differentiated participants by HIV status (Fig. 2h).

In summary, drastic metabolic alterations were observed in HEU infants and their mothers, with evidence of disrupted steroid, sulfur, sugar, bilirubin and bile acid metabolism. Many altered metabolites can be linked to the enzyme CYP2B6, which is known to both metabolize and be modulated by efavirenz[28]. Moreover, the metabolic impact was consistent with other known effects of efavirenz such as activating the pregnane X receptor, constitutive androstane receptor and neuronal receptors[29-31], and its adverse effects on lipid and sugar metabolism[32]. Notably, although increased morbidity and mortality and growth defects were previously reported in HEU infants born to mothers who did not receive efavirenz during pregnancy[6], in our study, the effect of efavirenz on the metabolomic profiles of mothers and infants dominated the differences between HEU and HUU mother-infant pairs. We present here the first MWAS analysis of global effects of efavirenz on human biochemistry. Besides the reported metabolites, many significant metabolite features in the dataset are still unidentified, an area for future investigations. Modern life is in constant chemical exposures, starting in utero. Our study demonstrates that chemical exposures can be tracked and assessed by untargeted metabolomic analysis, adding to the comprehensive understanding of human biochemistry in health and disease.

## Data Availability

All data produced in the present study are available upon reasonable request to the authors.

## Data and Code Availability

The serial dilution data are on Metabolomics Workbench (https://www.metabolomicsworkbench.org) under the study ID ST004665. All data analysis in this work is provided as Jupyter notebooks at https://github.com/shuzhao-li-lab/dark_metabolome.

## Acknowledgements

This work was in part supported by NIH grants R01 HD107793 (AW), R01 AI149746 (SL) and ARPA-H award D24AC00345. The content is solely the responsibility of the authors and does not necessarily represent the official views of the funders.

## Supplementary Information

**Supplemental Figure 1. Overview of HILIC-metabolomics data**.

a) PCA plot and b) clustermap showing the separation of HEU/HUU and maternal/cord samples in HILIC-metabolomics data.

**Supplemental Figure 2. Additional significant metabolites based on HIV exposure status, both infants and mothers**.

**Supplemental Figure 3. Many significant differential metabolites are mapped to a steroid metabolism network**. All shown metabolites are significant in the mothers’ data. Red indicates higher abundance in the HEU group.

a) Network of altered steroids due to HIV exposure in mothers.

b) Same network in a) but colored by fold change in the infants’ data.

**Supplemental Figure 4. Additional result on efavirenz exposure**.

a) Correlation plot highlighting the positive correlation of 8-hydroxyefavirenz between paired maternal and cord plasma samples.

b) Manhattan plot comparing the distribution of features correlated with Efavirenz versus features that are significantly different due to HIV exposure in maternal plasma samples along the retention-time axis.

c) Dot plot summarizing the pathway analysis results of efavirenz and HIV exposure status associated features in maternal and cord samples in RP-data, which have limited pathway coverage.

## Methods

### Cohort enrollment and sample collection

Enrollment took place in at the University of Witwatersrand between June and December 2017. The study was approved by the Ethics Committee of the Witwatersrand University and by the Colorado Multiple Institutions Review Board. The demographic characteristics of the study population, the results of the dietary questionnaire and of the maternal microbiome analysis were previously published[18]. The archived cord and maternal plasma samples collected at delivery from 123 mothers with HIV and 117 mothers without HIV are used here for metabolomics analysis.

### Metabolite and lipid extraction

Plasma samples were maintained on ice throughout the process. A pooled quality control (Pool_QC) sample was created by combining 10 µL aliquots from all samples in Batch 1. To prepare the internal standard (IS) solution for metabolomics, individual stock solutions of labeled metabolites were prepared in water: 2 mM L-Tyrosine-15N (Cambridge Isotope Laboratories, cat. no. NLM-590), 10 mM Uracil-15N2 (Cambridge Isotope Laboratories, cat. no. NLM-637), 20 mM L-Glutamic acid-13C5 (Cambridge Isotope Laboratories, cat. no. CLM-1800-H), 5 mM Caffeine-3-methyl-13C (Cambridge Isotope Laboratories, cat. no. CLM-728), 10 mM L-Methionine-13C5,15N (Cambridge Isotope Laboratories, cat. no. CNLM-759-H), and 1 M D-Glucose-13C6 (Cambridge Isotope Laboratories, cat. no. CLM-1396). A final IS solution was prepared by combining precise volumes of each stock solution with water to achieve desired concentrations, followed by vortexing for 30 seconds. IS for lipidomics was commercial LIPIDOMIX internal standard (Avanti Lipids, cat. No. 330707-1EA)

For metabolomics, extraction blanks (20 µL of water), Pool_QC samples (20 µL), commercial human plasma QC (Qstd; MilliporeSigma, cat. no. H4522; 20 µL), and NIST SRM 1950 plasma (MilliporeSigma, cat. no. NIST1950; 20 µL) were aliquoted into labeled tubes and processed identically to study samples, respectively. For extraction, 20 µL of plasma sample was mixed with 100 µL of ice-cold methanol and 4 µL of internal standard (IS) solution. Samples were vortexed for 30 s and incubated on ice for 10 min to precipitate proteins, followed by centrifugation at 21,000 × g and 4°C for 20 min. Subsequently, 80 µL of the supernatant was transferred to autosampler vials for LC–MS analysis.

Lipid extraction was performed using an MTBE-based biphasic protocol. An extraction buffer (30 mL MTBE supplemented with 1 mL LIPIDOMIX internal standard, Avanti Lipids, cat. No. 330707-1EA) and a lipid reconstitution buffer (8 mL methanol with 1 mL toluene) were prepared in advance and kept at 4°C. Plasma samples were maintained on ice throughout handling. Blank, Qstd, NIST SRM 1950, and Pool_QC samples were prepared as described for metabolomics and processed in parallel.

For extraction, 20 µL of plasma was mixed with 100 µL of ice-cold methanol and vortexed for 30 s. Next, 300 µL of cold extraction buffer was added and the mixture was vortexed, followed by addition of 100 µL water to induce phase separation. Samples were incubated on ice for 10 min and centrifuged at 21,000 × g and 4°C for 20 min. The upper organic phase (250 µL) was transferred to a fresh tube, dried in a vacuum concentrator at 30°C, and reconstituted in 80 µL of lipid reconstitution buffer. Reconstituted extracts were vortexed, centrifuged again to remove particulates, and 70 µL of the clarified supernatant was transferred to autosampler vials for LC– MS analysis.

### LC-MS and MS/MS Metabolomics analysis

Metabolomics and lipidomics analyses were performed using HILIC and reversed-phase (RP) C18 separations, respectively, coupled to a Thermo Scientific Orbitrap mass spectrometer, as described previously [33,34]. For HILIC metabolomics, an Accucore™ 150 Amide column set (guard, 10 × 2.1 mm, Thermo Fisher Scientific, cat. no. 16726-012105; analytical, 100 × 2.1 mm, cat. no. 16726-102130) was used. Mobile phases were prepared with 10 mM ammonium acetate (Fisher Scientific, cat. no. A11450) and 0.1% acetic acid (Fisher Scientific, cat. no. A11350) in acetonitrile/water. The gradient (mobile phase B, %) was programmed as follows: 0– 0.20 min, 0% B; 0.20–8.75 min, linear increase to 98% B; 8.75–10.00 min, held at 98% B; 10.00–15.00 min, returned to 0% B; and 15.00–20.00 min, held at 0% B for re-equilibration. The flow rate was 550 µL/min from 0–10.00 min, decreased to 100 µL/min at 15.00 min, restored to 550 µL/min at 17.00 min, and maintained at 550 µL/min until 20.00 min.

For RP lipidomics, Hypersil GOLD™ C18 columns (guard, 10 × 2.1 mm, Thermo Fisher Scientific, cat. no. 25003-012101; analytical, 50 × 2.1 mm, cat. no. 25003-052130) were used. Mobile phase A consisted of 60:40 acetonitrile:water (v/v), and mobile phase B consisted of 90:10 isopropanol:acetonitrile (v/v), each containing 10 mM ammonium formate (Fisher Scientific, cat. no. A11550) and 0.1% formic acid (Fisher Scientific, cat. no. A1171-AMP). The gradient (mobile phase B, %) was: 0–0.01 min, 15% B; 0.01–2.01 min, linear increase to 30% B; 2.01–2.51 min, linear increase to 48% B; 2.51–11.00 min, linear increase to 82% B; 11.00–11.50 min, linear increase to 99% B; 11.50–15.00 min, held at 99% B; 15.00–16.50 min, increased to 100% B; 16.50–17.50 min, returned to 15% B; and 17.50–20.00 min, held at 15% B for re-equilibration. The flow rate was maintained at 400 µL/min throughout.

MS data were acquired in both positive and negative ionization modes over m/z 66.7–1000.0 at a resolution of 60,000 (at m/z 200). Source parameters were optimized and included spray voltages of 3.5 kV (positive) and 2.5 kV (negative), a capillary temperature of 300°C, and a heater temperature of 425°C.

Data dependent MS2 (DDA) was applied to subpool samples over mass ranges (66.7–1000.0 m/z) using the following settings: for Full MS, resolution = 60,000; AGC target = 3e6; Maximum IT = 50 ms; for dd-MS^2^: resolution = 30,000; AGC target = 1e5; Maximum IT = 50 ms; Isolation width = 1.0 m/z; stepped normalised collision energies = 20, 30, 40%.

### Data acquisition and preprocessing, annotation

Metabolomics data from four acquisition modes (HILIC ESI+, HILIC ESI-, RP ESI+, RP ESI-) were processed using asari version 1.14[33] and the PCPFM pipeline version 1.1[35]. Processed feature tables were blank-masked, outlier-filtered, normalized by TIC, log2 transformed, and batch-corrected before downstream statistical analyses. Batch-corrected feature tables were imported into Python (v3.11) using pandas. Sample metadata were merged to create comprehensive sample–feature mappings, including participant status (HEU vs. HUU) and sample type (maternal or cord plasma). Annotations at multiple confidence levels (MS/MS-based L2–L1a) were indexed for downstream analysis.

### Statistical analysis

Statistical analyses were performed individually for each metabolic mode. First, to obtain a global overview of all samples, principal component analysis (PCA) was performed in R 4.5.1 using the prcomp function from the stats package, with centering and scaling set to TRUE. Next, paired t-tests between matched mother-baby pairs was applied using the col_t_welch function from matrixTests to account for heteroscedasticity. Multiple-testing correction was computed across all analyses using the false discovery rate (FDR, Benjamini–Hochberg). Now within

mothers and cords separately, Welch’s t-tests (row_t_welch, matrixTests) and fold-changes were calculated between HEU vs HUU groups.

Correlation networks between drug-related features and other metabolites were computed using pairwise Pearson correlations within group-specific subsets (HEU or HUU). Observed correlation distributions were compared to null distributions generated through permutation testing (1,000 iterations), yielding empirical z-scores and permutation-based p-values to assess enrichment beyond random expectation. Significant associations were visualized in enrichment histograms with annotated overlays.

### Data visualization

For PCA visualization, a plot of the first two principal components were genereated per metabolic mode, colored by HIV exposure status and sample origin (mathernal vs cord). 95% confidence ellipses were overlapid based on the covariance structure of each group to highlight clustering. To visualize the similarity of significant features across all samples, a heatmap of metabolic features with a BH-adjusted p-value < 0.05 from the paired t-tests was plotted using the ComplexHeatmap R package. Batch, origin (mother vs cord), and status (HIV exposure status) were annotated on the heatmap columns. Feature intensities were z-score standardized using the scale function in R.

Volcano plots were generated using ggplot2 to visualize statistical significance (−log_10_ p-values) versus effect size (log_2_ fold change). Points were color-coded by significance category: both fold-change and p-value significant, or non-significant. Annotations for efavirenz and its metabolites (Levels 1a–2) were highlighted on the volcano plot. Plots were generated separately for maternal and cord plasma. For metabolites of particular interest (e.g., drug-related compounds and key metabolic markers), boxplots were generated to display group-wise distributions. These plots utilized alpha-transparency overlays to indicate replicate density and included adjusted p-values (FDR) to support feature-level interpretation.

Plots were generated with ggplot2 and ggrepel for clear annotation and high-resolution output. Colors were standardized (HEU: #D41159, red; HUU: #1A85FF, blue) across all plots for consistency. Boxplots, PCA, volcano, and enrichment plots were archived to a reproducible directory structure for integration with manuscript figures. All analyses were scripted in R (tidyverse, ggplot2, ggrepel, matrixTests, ComplexHeatmap), and all notebooks were version-controlled to ensure reproducibility.

### Pathway and Enrichment Analysis

Significant features were formatted for pathway enrichment analysis using Mummichog version 2.7. Separate input tables were generated for maternal and cord samples per ionization mode, including m/z, retention time, raw p-value, and test statistic. Mummichog was executed to identify pathways disproportionately associated with significant metabolites, and pathway tables were sorted by overlap size and adjusted significance for downstream visualization. To visualize the Mummichog results for efavirenz-correlated and differentially abundant features due to HIV exposure in cord and maternal samples separately, a dot plot was created using ggplot2. The size of each dot corresponded to the -log10 p-value, and the dot was either filled (p-value < 0.05) or empty (p-value > 0.05).

## References

1. Al-Haddad, B.J.S., et al., Long-term Risk of Neuropsychiatric Disease After Exposure to Infection In Utero. JAMA Psychiatry, 2019. 76(6): p. 594–602.

2. Cohn, B.A., et al., DDT Exposure in Utero and Breast Cancer. J Clin Endocrinol Metab, 2015. 100(8): p. 2865–72.

3. Jacobson, J.L., S.W. Jacobson, and H.E. Humphrey, Effects of in utero exposure to polychlorinated biphenyls and related contaminants on cognitive functioning in young children. J Pediatr, 1990. 116(1): p. 38–45.

4. Almond, D., Is the 1918 influenza pandemic over? Long-term effects of in utero influenza exposure in the post-1940 US population. Journal of political Economy, 2006. 114(4): p. 672–712.

5. Lin, M.-J. and E.M. Liu, Does in utero exposure to illness matter? The 1918 influenza epidemic in Taiwan as a natural experiment. Journal of health economics, 2014. 37: p. 152–163.

6. Evans, C., C.E. Jones, and A.J. Prendergast, HIV-exposed, uninfected infants: new global challenges in the era of paediatric HIV elimination. Lancet Infect Dis, 2016. 16(6): p. e92–e107.

7. Slogrove, A.L., L.F. Johnson, and K.M. Powis, Population-level Mortality Associated with HIV Exposure in HIV-uninfected Infants in Botswana and South Africa: A Model-based Evaluation. J Trop Pediatr, 2019. 65(4): p. 373–379.

8. Wedderburn, C.J., et al., Early neurodevelopment of HIV-exposed uninfected children in the era of antiretroviral therapy: a systematic review and meta-analysis. Lancet Child Adolesc Health, 2022. 6(6): p. 393–408.

9. Bulterys, M.A., et al., Neurodevelopment among children exposed to HIV and uninfected in sub-Saharan Africa. J Int AIDS Soc, 2023. 26 Suppl 4(Suppl 4): p. e26159.

10. Bowen, T.J., et al., Simultaneously discovering the fate and biochemical effects of pharmaceuticals through untargeted metabolomics. Nature Communications, 2023. 14(1): p. 4653.

11. Fischer, S.T., et al., Low-level maternal exposure to nicotine associates with significant metabolic perturbations in second-trimester amniotic fluid. Environ Int, 2017. 107: p. 227–234.

12. Li, S., et al., Understanding mixed environmental exposures using metabolomics via a hierarchical community network model in a cohort of California women in 1960’s. Reprod Toxicol, 2020. 92: p. 57–65.

13. Miller, G.W. and B.E. Consortium, Integrating exposomics into biomedicine. Science, 2025. 388(6745): p. 356–358.

14. Moutloatse, G.P., et al., Metabolic risks at birth of neonates exposed in utero to HIV-antiretroviral therapy relative to unexposed neonates: an NMR metabolomics study of cord blood. Metabolomics, 2016. 12(11): p. 175.

15. Moutloatse, G.P., et al., Metabolic risks of neonates at birth following in utero exposure to HIV-ART: the amino acid profile of cord blood. Metabolomics, 2017. 13(8): p. 89.

16. Kaur, S.U., et al., Plasma metabolomic study in perinatally HIV-infected children using 1H NMR spectroscopy reveals perturbed metabolites that sustain during therapy. PLoS One, 2020. 15(8): p. e0238316.

17. Schoeman, J.C., et al., Fetal Metabolic Stress Disrupts Immune Homeostasis and Induces Proinflammatory Responses in Human Immunodeficiency Virus Type 1- and Combination Antiretroviral Therapy-Exposed Infants. J Infect Dis, 2017. 216(4): p. 436–446.

18. Jackson Conner, L., et al., Evolution of the Gut Microbiome in HIV-Exposed Uninfected and Unexposed Infants during the First Year of Life. mBio, 2022. 13(5): p. e01229–22.

19. Isvoran, A., et al., Pharmacogenetics of human sulfotransferases and impact of amino acid exchange on Phase II drug metabolism. Drug Discov Today, 2022. 27(11): p. 103349.

20. Cetin, I., G. Alvino, and M. Cardellicchio, Long chain fatty acids and dietary fats in fetal nutrition. J Physiol, 2009. 587(Pt 14): p. 3441–51.

21. Solano, M.E. and P.C. Arck, Steroids, Pregnancy and Fetal Development. Front Immunol, 2019. 10: p. 3017.

22. de Aguiar Vallim, T.Q., E.J. Tarling, and P.A. Edwards, Pleiotropic roles of bile acids in metabolism. Cell Metab, 2013. 17(5): p. 657–69.

23. Fiore, A. and P.J. Murray, Tryptophan and indole metabolism in immune regulation. Curr Opin Immunol, 2021. 70: p. 7–14.

24. Xue, C., et al., Tryptophan metabolism in health and disease. Cell Metab, 2023. 35(8): p. 1304–1326.

25. Li, S., et al., Predicting network activity from high throughput metabolomics. PLoS Comput Biol, 2013. 9(7): p. e1003123.

26. Cressey, T.R., et al., Efavirenz pharmacokinetics during the third trimester of pregnancy and postpartum. J Acquir Immune Defic Syndr, 2012. 59(3): p. 245–52.

27. Kreitchmann, R., et al., Efavirenz pharmacokinetics during pregnancy and infant washout. Antivir Ther, 2019. 24(2): p. 95–103.

28. Wang, P.-F., A. Neiner, and E.D. Kharasch, Efavirenz metabolism: influence of polymorphic CYP2B6 variants and stereochemistry. Drug Metabolism and Disposition, 2019. 47(10): p. 1195–1205.

29. Gatch, M.B., et al., The HIV antiretroviral drug efavirenz has LSD-like properties. Neuropsychopharmacology, 2013. 38(12): p. 2373–84.

30. Sharma, D., et al., Agonism of human pregnane X receptor by rilpivirine and etravirine: comparison with first generation non-nucleoside reverse transcriptase inhibitors. Biochem Pharmacol, 2013. 85(11): p. 1700–11.

31. Sharma, D., et al., Differential activation of human constitutive androstane receptor and its SV23 and SV24 splice variants by rilpivirine and etravirine. Br J Pharmacol, 2015. 172(5): p. 1263–76.

32. Haugaard, S.B., Toxic metabolic syndrome associated with HAART. Expert Opin Drug Metab Toxicol, 2006. 2(3): p. 429–45.

33. Li, S., et al., Trackable and scalable LC-MS metabolomics data processing using asari. Nature Communications, 2023. 14(1): p. 4113.

34. Siddiqa, A., et al., A pilot metabolomic study of drug interaction with the immune response to seasonal influenza vaccination. npj Vaccines, 2023. 8(1): p. 92.

35. Mitchell, J.M., et al., Common data models to streamline metabolomics processing and annotation, and implementation in a Python pipeline. PLOS Computational Biology, 2024. 20(6): p. e1011912.

