## Supplementary figures and images for "Distinct biochemical phenotypes of HIV exposed infants driven by antiviral medication" (from another version of this preprint: version 1, 2026-02-02)

### Supplemental Figures 1-4

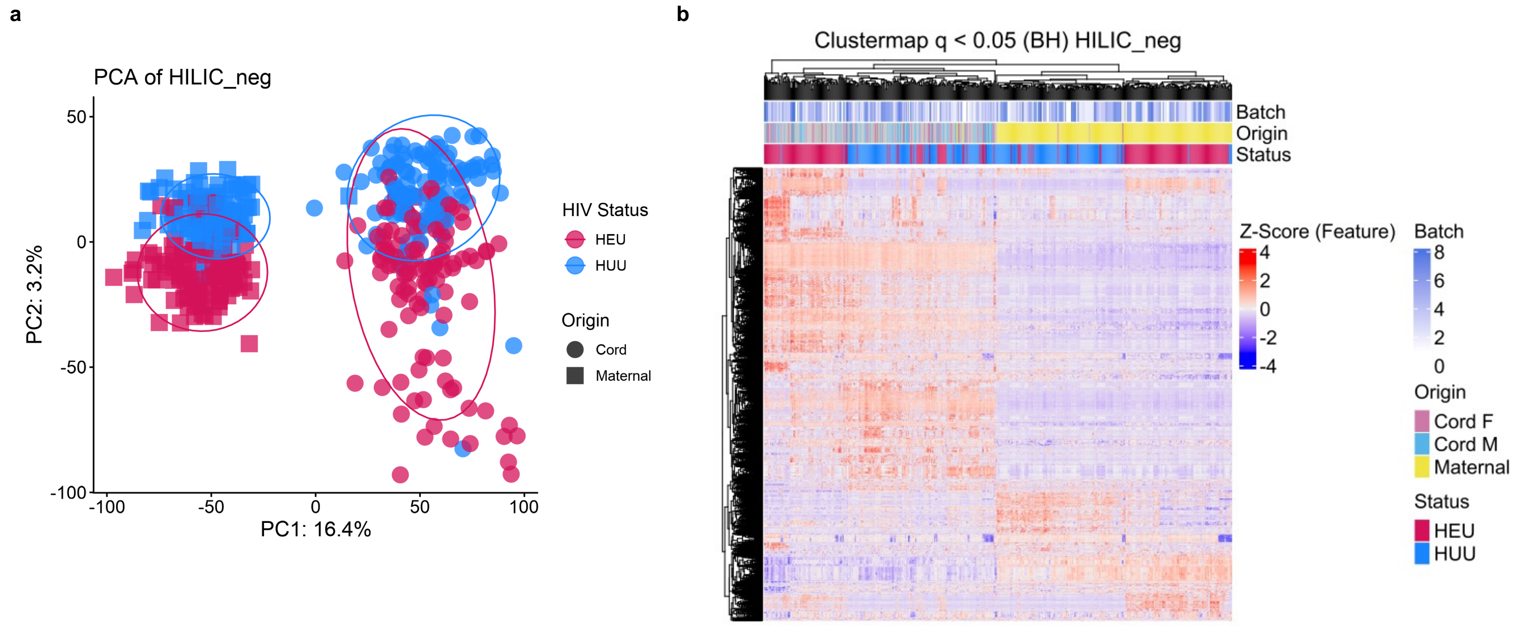


#### Supplementary Figure 1

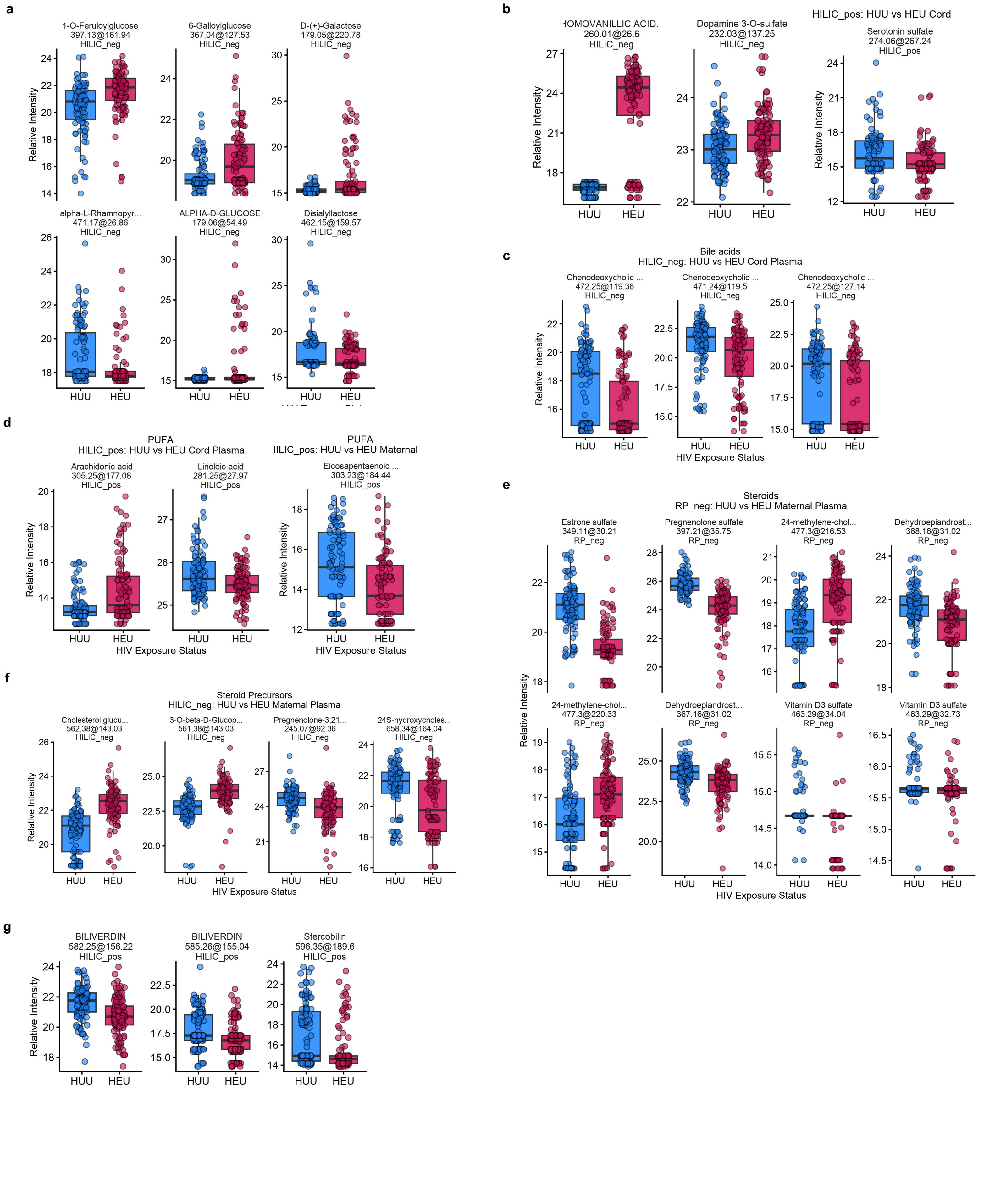


Supplementary Figure 2

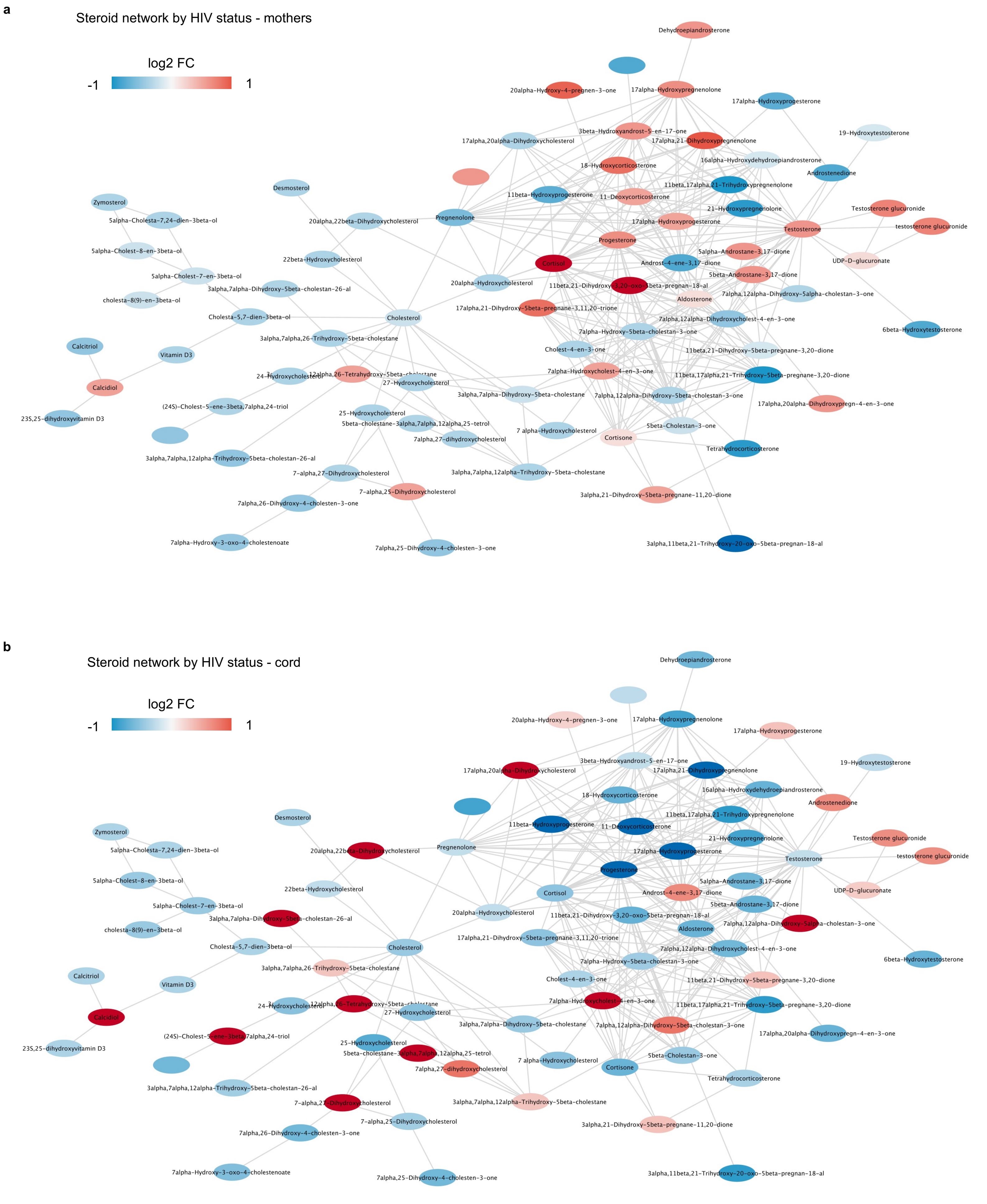


Supplementary Figure 3

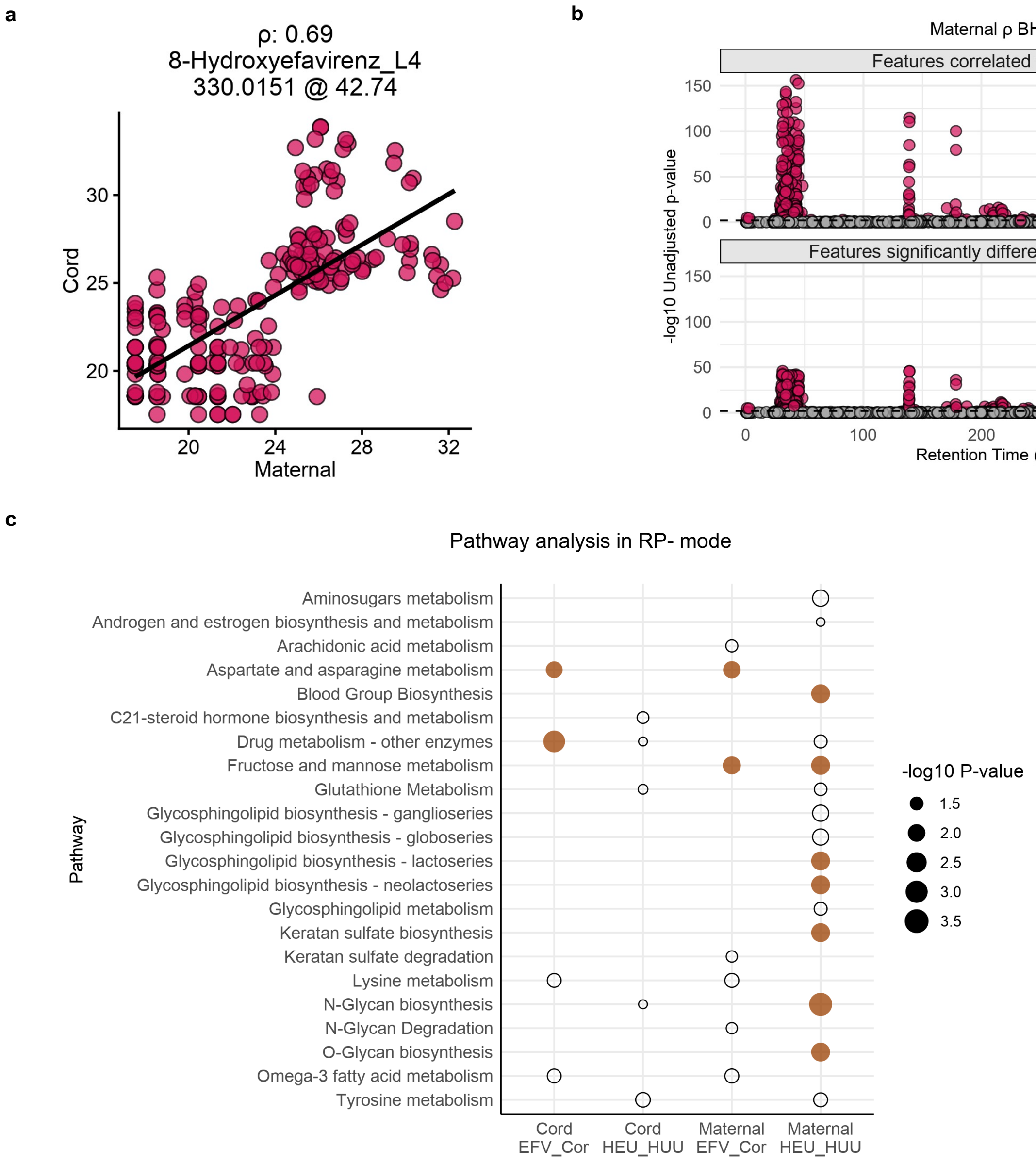


Supplementary Figure 4
